# The economic effects of kidney failure treatment on the household welfare of patients on Dialysis in Buea and Bamenda–Cameroon

**DOI:** 10.1101/2023.12.05.23299509

**Authors:** Therence Nwana Dingana, Stewart Ndutard Ngasa, Neh Chang Ngasa, Leo Fosso Fozeu, Fuein V. Kum, Njong M. Aloysius

**Affiliations:** Medical Research and Careers Organisation; Tubah District Hospital, Regional Delegation of Public Health, Northwest Region, Cameroon; The Spinney, Elysium Healthcare, Manchester, United Kingdom; Ndop District Hospital, Regional Delegation of Public Health, Northwest Region, Cameroon; Faculty of Health Sciences, University of Buea

**Keywords:** kidney failure, dialysis, healthcare cost, household welfare

## Abstract

**Background:** Noncommunicable diseases, such as kidney failure, diabetes, and cancer, are among the leading causes of death worldwide. There is a sharp increase in the incidence and prevalence of patients with kidney failure requiring replacement therapy. This has led to a very high cost, especially in resource-limited settings like Cameroon. The aim of this study is to determine the effects of direct and indirect costs of kidney failure treatment on their household income.

**Methods:** A descriptive cross-sectional study design was used. Primary data was collected using a self-administered pre-tested questionnaire for the economic impact of chronic disease. For bivariate analysis, we used the Cochran-Mantel-Haenszel test to obtain crude Odd Ratios (OR) of factors associated with household welfare. Multivariate logistic regression, the OLS model was used to identify independent associations between kidney failure treatment and household welfare. This was presented as adjusted odd ratios along with their p-values. A p-value of <0.05 was used as a cut-off for statistical significance.

**Results:** The mean age of participants was 44.6±15.5 years; most patients (83(62.4%)) were married. Seventy-nine (59.4%) were unemployed, and eighty-one (60.9%) had no financial support. Their annual household expenditure ranged from 300,000FCFA to 3,360,000FCFA, with a mean and standard deviation of 1,547,729FCFA and 781,882FCFA, respectively. The yearly total cost of kidney failure treatment ranged from 520,000FCFA to 10,000,000FCFA with a mean and standard deviation of 2,137,556FCFA and 1,541,163FCFA, respectively. The cost of consultation and laboratory tests had negative regression coefficients (P=0.001 and <0.001 respectively).

**Conclusion:** kidney failure has a significant negative effect on the household welfare of patients on dialysis. Kidney disease screening and prevention programs are necessary to reduce the number of persons in need of hemodialysis. Health insurance schemes and universal health coverage should target patients on hemodialysis.

## Background

Globally, disease burden is rising, ranging from noncommunicable diseases, malnutrition, neglected tropical diseases, and infectious disease afflictions^1^. However, global attention has focused on infectious diseases like HIV, tuberculosis, and the COVID-19 pandemic. Noncommunicable diseases, such as heart disease, cancer, kidney failure, chronic respiratory disease, and diabetes, contribute a lot to global health miseries and are the leading causes of death worldwide and, therefore, represent an emerging international health threat. Deaths from noncommunicable diseases are far more than all communicable diseases^2^. Noncommunicable diseases end the lives of 41 million people each year, corresponding to over 7 out of 10 deaths globally.

Kidney failure is one of the main global noncommunicable diseases lacking proper attention, especially in resource-limited settings. There is a sharp rise in the incidence and prevalence of patients with kidney failure requiring dialysis, whose cost is usually very high^3^. The International Society of Nephrology projected that in 2030, 14.5 million people will have kidney failure and need treatment. However, only 5.4 million will actually receive it due to economic, social, and political factors^4^.

The handiness of dialysis and kidney transplantation for treating kidney failure has been one of medicine’s greatest successes in past decades. It has been accessible in developed countries for over 50 years, with an increasing number of patients being treated^5,6^. The use of dialysis varies regionally due to differences in population demographics, the prevalence of End-Stage Renal Disease (ESRD) and, most specifically, access to and provision for Renal Replacement Therapy (RRT)^7,8^. Treating renal failure is disproportionately costly compared to other medical conditions and constitutes a heavy burden on communities and households worldwide^6^ Available data on the cost of RRT in low-income nations, especially in sub-Saharan Africa (SSA) is scarce, as opposed to high and middle-income countries^6,9,10^. Since 2010, the USA spent about $28 billion yearly for ESRD-related medical expenses^11^. In the UK, the management cost for ESRD was 1–2% of the funds of the National Health Service for patients who make up only 0.05% of the population^11,6^. Dialysis is an example of a robust single-payer dominant system in the United States, unlike most low-income countries like Cameroon, where payments are mostly “out of pocket”^12^; where households are required to do ‘pay as they go’. Consequently, poor households quickly face catastrophic health expenditures, become financially drained, income depleted, and unmanageable in the context of kidney failure^13^.

Treatment of chronic kidney disease is very precise, comprising medical consultations, laboratory investigations, dialysis/kidney transplant, drug therapy, and lifestyle adjustments^14^. All these are very costly and can be challenging for the patients to cope with. In most developed countries like the United States, patients receiving dialysis are insured by Medicare, a robust single-payer dominant system^15^

Dialysis is comparatively more expensive for poorer than wealthier developing countries and may not be cost-effective for poor countries such as Cameroon^16,17^. In Cameroon, just 5.1% ($ 1.3 billion) of the state budget is assigned to healthcare. With other pressing health concerns such as high maternal and infant mortality and HIV/AIDS, haemodialysis becomes a serious economic burden on the healthcare sector and therefore almost all the costs are left for the patients to bear. Poor households quickly become financially drained while wealthy households comfortably survive. This shows a significant disparity in the economic impact on the households. Lack of resources, limited access or high cost of treatment results in under-diagnosis of kidney failure, and even those diagnosed end up receiving less care than required (or resort to ineffective measures like prayers or traditional medicine) leading to pitiable health outcome ^18^

The current socio-political crisis plaguing the English-speaking part of Cameroon has brought more challenges to these patients, in terms of abandonments of settlements, death of breadwinners; cost of transportation, lockdown days, and travel risks etc., all of these will increase the socioeconomic burden of kidney failure. Available knowledge on the economic effects of kidney treatment on household welfare in this area lacking. We, therefore, resorted to evaluating the economic effects of kidney failure treatment on the household welfare of patients in the dialysis centers of Buea and Bamenda – Cameroon. This evaluation consisted of evaluating the direct and indirect cost of kidney failure treatment.

## Methods

### Study design, setting and participants

Cameroon’s South West and North West regions, with a population of about 3.52 million, are faced with a dual emergency; the socio-political crisis, which turned violent in November 2017, and recently the COVID-19 disease. These regions have approximately 400 patients requiring dialysis and have two hemodialysis centers, the Buea Regional Hospital and the Bamenda Regional Hospital.

The financial burden of kidney failure affects many levels; the government, firms, society, and households. This study was focused on the economic burden at the household level. The content of the economic effect in this study is composed of the direct and indirect costs of kidney failure. The monthly expenses were analyzed and projected for one year (2022). Dialysis patients for acute kidney injury and other causes like hyperkalemia requiring just a few dialysis sessions were excluded. Patients who initiated dialysis less than a month ago were exempted from the study.

### Sampling and data collection

We used the descriptive cross-sectional study design and data collection started in May 2023. Primary data was collected using a self-administered pre-tested questionnaire for the economic impact of chronic disease^19^. The questionnaire has mainly closed-ended questions and divided into three main sections. Section A is general information and household socio-demography and household expenditure, section B is the direct cost of kidney failure, and section C is the indirect cost of kidney failure. All collected data was anonymised.

#### Description of main variables

The study assessed several economic factors related to kidney failure treatment. Household expenditure (Y) was quantified in CFA francs, which involved approximating the patients’ monthly spending. To determine the average annual household expenditure, the average monthly spending was multiplied by 12.

Direct cost (X1) represented the total annual cost of treating kidney failure, encompassing expenses such as consultation fees, laboratory tests, medication costs, medical devices, self-medications, other treatments like special diets, dialysis costs, and additional expenses related to dialysis. These costs were projected for a one-year duration.

Indirect cost (X2) referred to the average annual expenses associated with kidney failure treatment but not directly linked to medical procedures. This included transport expenses, parking fees, accommodation costs, caregiver fees, informal caregiver expenses, the cost of accompanying persons, caregiver accommodation, and other related costs. Like direct costs, these expenses were projected over a one-year period.

To examine the impact of these factors on household expenditure and, consequently, household welfare, the study employed an ordinary least square (OLS) approach due to the continuous nature of the dependent variables. OLS estimators involve linear functions of the household expenditure (Y) values, connected through weights that are a non-linear function of the direct and indirect cost values (X1 and X2). The analysis was conducted using the software SPSS, which generated values for the unknowns (β0, β1, β 2) in the linear model equation.

The research employed the Ordinary Least Squares regression model to investigate the relationship between the direct and indirect costs of kidney failure treatment and household welfare. Household welfare was considered the dependent variable, while direct and indirect costs were treated as independent variables. This relationship can be summarized by the following function:

Household welfare (HW) = F (direct cost, indirect cost);

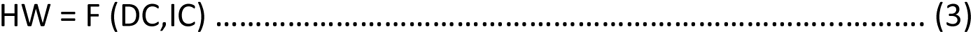

In our study, we measured household welfare by measuring household consumption in terms of household expenditure as specified by World Bank (2000).

Using the multiple regression models, we transformed the function into a multiple regression equation for empirical verification as follows:

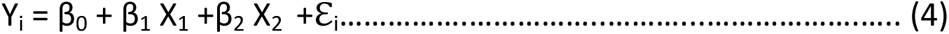

Where:

Yi= HE (household expenditure), the dependent variable and a measure of household welfare X1= Direct cost (DC) and X2= Indirect cost (IC) are the independent variables ℇi, = Error term, which constitutes other predictors of household welfare not considered in our model.

Our data set will then help us to get estimates for β0, β1, and β2,

### Sample size calculation

We determined the sample size for this work based on Taro Yamane’s approach to finite populations^20^

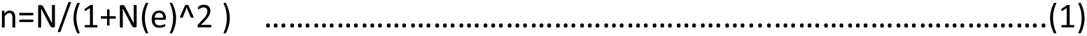

Where,

n = the minimum sample size

N = the finite population out of which the sample was taken

e = the acceptable sampling error (or limit of tolerable error)

The total number of patients permanently on dialysis in Buea is 95, and Bamenda is 71 giving a total of 166, setting the significance level at 0.05 or 5%. Therefore, the minimum sample size (n) was calculated as

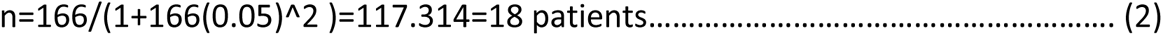

We used the stratified sampling technique to recruit the participants. These participants were grouped as men, women, and children. One hundred and thirty-three (133) participants took part in the study.

### Statistical methods and data analysis

Data were entered into excel spreadsheet and analysed using Stata version 14 statistical software. Results were presented as means and standard deviation (SD) for continuous variables and frequencies and percentages for categorical variables. At bivariate analysis, we used the Cochran-Mantel-Haenszel test to obtain crudes Odd Ratios (OR) of factors associated with household welfare. Multivariate logistic regression was used to identify independent associations with kidney failure treatment and household welfare. This was presented as adjusted odd ratios along with their p-values. A p-value of <0.05 was used as cut off for statistical significance.

## Results

### Sociodemographic characteristics of participants

The patient ages exhibited a normal distribution and varied from 13 to 80 years, with an average age of 44.6±15.5 years. The majority of patients fell in the age range of 30 to 60 years, making up 63.9% of the sample (refer to Table 1). Out of the 133 participants, 80 (60.2%) were male, while 53 (39.8%) were female. In terms of marital status, the majority of patients (83 or 62.4%) were married, 38 (28.6%) were single, 10 (7.5%) were widows or widowers, and 2 (1.5%) were divorced. Regarding employment, 79 (59.4%) of the patients were unemployed, while 54 (41%) were employed. Educational backgrounds varied, with 55 (41.4%) having attended secondary education, 38 (28.6%) having completed primary education, 39 (29.3%) having tertiary education, and one patient having no formal education.

**Table 1:** Sociodemographic characteristics of Participants.

| Variable | Number | Percentage |
| --- | --- | --- |
| **Gender** |  |  |
| Male | 80 | 60.2 |
| Female | 53 | 39.8 |
| **Marital Status** |  |  |
| Married | 83 | 62.4 |
| Single | 38 | 28.6 |
| Divorced | 2 | 1.5 |
| Widow | 10 | 7.5 |
| **Employment status** |  |  |
| Employed | 54 | 40.6 |
| Unemployed | 79 | 59.4 |
| **Level of education** |  |  |
| None | 1 | 0.8 |
| Primary | 38 | 28.6 |
| Secondary | 55 | 41.4 |
| Tertiary | 39 | 29.3 |
| **Who pays bills** |  |  |
| Self | 81 | 60.9 |
| Parents | 24 | 18.0 |
| Insurance | 0 | 0.0 |
| Spouse | 7 | 5.3 |
| Children | 18 | 13.5 |
| Others | 3 | 2.3 |
| **Comorbidities** |  |  |
| None | 39 | 29.3 |
| Heart disease | 3 | 2.3 |
| Diabetes | 12 | 9.0 |
| Hypertension | 86 | 64.7 |
| Other | 3 | 2.3 |
| **Variable** | **Mean ± SD** | **Median (IQR)** |
| Age (years)* | 44.6 ± 15.5 | 43 (34 – 55.5) |
| Duration on dialysis (months) | 28.9 ± 36.4 | 12 (7 – 33) |
| House hold density | 5.7 ± 2.8 | 6 (4 – 7) |
| Duration in hospital per session (hours) | 17.5 ± 11.1 | 15 (7.5 – 24) |
| Time spent travelling (mins) | 57.2 ± 45.8 | 45 (30 – 90) |
| Annual household income | 1,547,729 ± 781,882 | 1,500,000(1.5 - 2.1M) |

Only a small proportion (39 or 29.3%) of patients had no comorbidities. Among the 94 (70.6%) patients with comorbidities, 86 (64.7%) had chronic hypertension, 12 (9.0%) had diabetes, 3 (2.3%) had heart disease, and 3 (2.3%) had other chronic illnesses, such as liver failure. Ten (7.5%) of the patients had both hypertension and diabetes. Household density ranged from 2 to 15 individuals per household, with an average of 5.7±2.8 persons per household. The duration of time the patients had been on dialysis ranged from 1 month to 168 months (equivalent to 14 years), with an average duration of 28.9±36.4 months (approximately 2.4±3.0 years). The time spent in the hospital for each dialysis session varied widely, spanning from 5 to 48 hours, with an average duration of 17.5±11.1 hours. The time patients needed to travel to the hospital ranged from 0 minutes (for those residing in the hospital) to 240 minutes (4 hours), with a mean travel time of 57.2±45.8 minutes.

None of the patients had their medical bills covered by health insurance, as illustrated in Figure 1. The majority (60.9%) were responsible for paying their own bills, while 24 (18.0%) had their bills covered by their parents. Additionally, 18 (13.5%) of the patients had their children covering the bills, 7 (5.3%) had their spouses paying the bills, and 3 (2.3%) had their bills covered through alternative means, such as by their siblings (refer to Figure 1).

#### Total household expenditures

The annual household expenditure ranged from 300,000FCFA to 3,360,000FCFA. The mean, median, and standard deviation of annual household expenditures were 1.547.729FCFA, 1.500.000FCFA, and 781.882FCFA, respectively.

None of the patients had their bills covered by health insurance. Eighty-one (60.9%) were paying their bills by themselves, and 24(18.0%) had their bills paid by their parents, while children paid the bills of 18(13.5%) of the patients, 7(5.3%) of the participants reported that their bills were paid by their spouses and 3(2.3%) of the participants’ bills were paid by other means like the siblings.

### Total household expenditures

The annual household expenditure ranged from 300,000FCFA to 3,360,000FCFA. The mean, median, and standard deviation of annual household expenditures were 1.547.729FCFA, 1.500.000FCFA, and 781.882FCFA, respectively.

### Direct cost of kidney failure

The annual direct cost of kidney failure treatment ranged from 520,000FCFA to 7,160,000FCFA with a mean and standard deviation of 1,648,176FCFA and 1,213,777FCFA, respectively.

### Indirect cost of kidney failure

The annual indirect cost of kidney failure treatment ranged from 00FCFA to 3,596,000FCFA with a mean and standard deviation of 489,380FCFA and 620,519FCFA, respectively (Table 2). The annual total cost of kidney failure treatment ranged from 520,000FCFA to 10,000,000FCFA with a mean and standard deviation of 2,137,556FCFA ($3, 320) and 1,541,163FCFA. To estimate the effect of kidney failure treatment on household welfare, we used household consumption measured in terms of household expenditure. The annual direct and indirect cost of kidney failure treatment both have negative regression coefficients of −0.022 and −0.147 respectively, (p=0.814 and p=0.114, respectively). The net effect of spending on kidney failure treatment had a regression coefficient of −0.119 (p=0.172) on household expenditure.

**Table 2:**
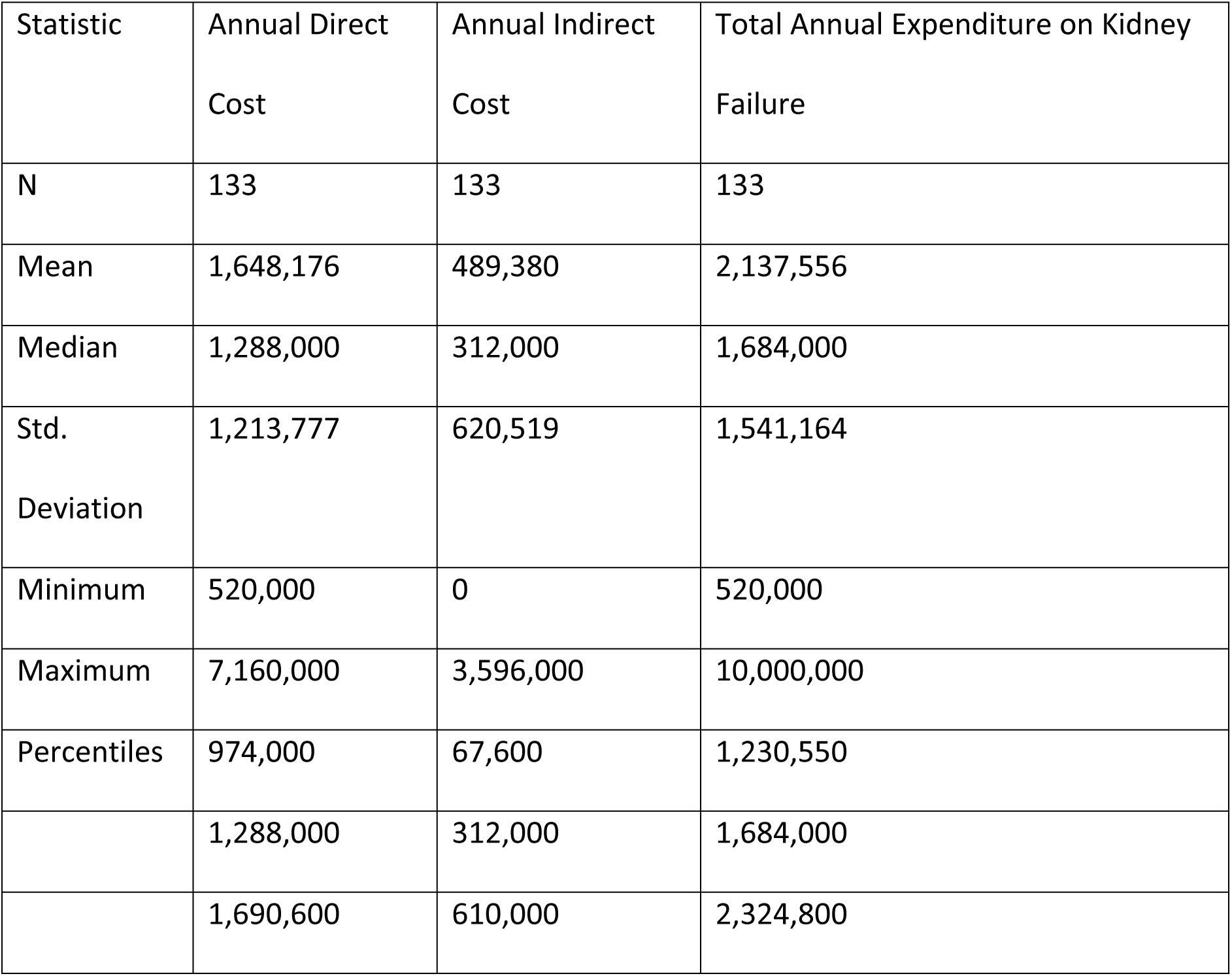
Annual Direct, Annual Indirect, and Total annual expenditure on kidney failure treatment.

| Statistic | Annual Direct Cost | Annual Indirect Cost | Total Annual Expenditure on Kidney Failure |
| --- | --- | --- | --- |
| N | 133 | 133 | 133 |
| Mean | 1,648,176 | 489,380 | 2,137,556 |
| Median | 1,288,000 | 312,000 | 1,684,000 |
| Std. Deviation | 1,213,777 | 620,519 | 1,541,164 |
| Minimum | 520,000 | 0 | 520,000 |
| Maximum | 7,160,000 | 3,596,000 | 10,000,000 |
| Percentiles | 974,000 | 67,600 | 1,230,550 |
|  | 1,288,000 | 312,000 | 1,684,000 |
|  | 1,690,600 | 610,000 | 2,324,800 |

From equation (2) above,

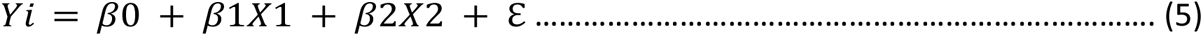

This implies,

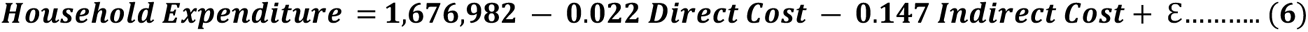

### Effects of the direct cost components of kidney failure treatment on the household expenditure of dialysis patients

Evaluating the regressions of each direct cost component reveals that the cost of consultation, laboratory tests, other treatment costs (nutrition), and cost of medical devices had negative regression coefficients of −0.293, −0.358, −0.005, and −0.07, respectively as on Table 4. These were statistically significant only for the consultation and laboratory test fees, with p-values of 0.001 and <0.001, respectively.

Drugs cost and other drugs cost (auto-medication) had positive regression coefficients of 0.189 and 0.327, respectively, with p-values of 0.028 and <0.001, respectively.

### Effects of indirect cost of kidney failure treatment on the household expenditure of dialysis patients

The regression analysis of each indirect cost component showed that the cost of transportation, caregiver accommodation, informal caregiver cost, and other costs on caregivers had negative regression coefficients of −0.131, −0.055, −0.059, and −0.094, respectively as on table 5. However, none of these were statistically significant.

Parking cost, accommodation cost, and caregiver cost caregiver transport cost all had positive regression coefficients of 0.164, 0.012, 0.004, and 0.127, respectively, however, none of them were statistically significant.

## Discussion

### Direct cost of kidney failure

The annual direct cost of kidney failure treatment ranged from 520,000FCFA to 7,160,000FCFA with a mean and standard deviation of 1,648,176FCFA and 1,213,777FCFA, respectively as on Table 2. This was way too low compared to the 7,678,553FCFA obtained by Halle and collaborators in tertiary hospitals in Cameroon^5^; this is simply because their study was not focused on household expenditure. They had a 30% cost covered by out-of-pocket while we had 100% out-of-pocket payments. Computing 30% of their value (7 678 553FCFA) gives 2,303,500, which is very close to the figure obtained in our study.

### Indirect cost of kidney failure

The annual indirect cost of kidney failure treatment ranged from 00FCFA to 3,596,000FCFA with a mean and standard deviation of 489,380FCFA and 620,519FCFA, respectively as on Table 2. This was very similar to the mean value of 530,118FCFA obtained by Halle and collaborators in tertiary hospitals in Cameroon^5^. However, their study was not a household-based study like ours.

### Total cost of kidney failure

The annual total cost of kidney failure treatment ranged from 520,000FCFA to 10,000,000FCFA with a mean and standard deviation of 2,137,556FCFA ($3 320) and 1,541,163FCFA, respectively as on Table 2. Our annual cost was approximately $ 3 320, with out-of-pocket payments being the main payment method. The annual cost of dialysis has been estimated at $ 87 500 in the USA^21^, 5,736 in India^22^, between $ 22 000–55 000 in Nigeria^23^, $ 27 440 in Tanzania^24^. One of the reasons for our lower cost is that we did not include the staff and building costs as has been done in other studies and all the costs considered are household expenditures and not general costs like in the other studies.

### Effects of kidney failure treatment on the household welfare of dialysis patients

We used household consumption measured in terms of household expenditure as a metric for household welfare. The annual direct and indirect cost of kidney failure treatment both have negative regression coefficients of −0.022 and −0.147 respectively, suggesting that as the direct and indirect cost of kidney failure treatment increase, the household expenditure decreases. This is very logical because the patients incur these health costs and are left with little to spend on other household needs and hence a negative impact on their household welfare. These coefficients, however, are close to zero, indicating a weak effect. Furthermore, these effects were not statistically significant (*p* = 0.814 and *p* = 0.114, respectively). The net effect of spending on kidney failure treatment had a regression coefficient of −0.119 with household expenditure, but this effect was not also statistically significant (*p* = 0.172) as on Table 3.

**Table 3:** OLS analysis for the direct, indirect, and total annual costs associated with average annual household expenditure.

| | Variables | $\beta_0$ | $\beta$ coef | p-value | R squared | |
| --- | --- | --- | --- | --- | --- | --- |
|  |  | 1,676,982 |  |  |  |  |
|  | Annual Direct cost |  | -0.022 | 0.814 | 0.024 |  |
|  | Annual Indirect cost |  | -0.147 | 0.113 |  |  |
|  | Total annual cost |  | -0.119 | 0.172 |  |  |

### Effects of the direct cost components of kidney failure treatment on the household expenditure of dialysis patients

Evaluating the regressions of each direct cost component reveals that the cost of consultation, laboratory tests, other treatment costs (nutrition), and cost of medical devices had negative regression coefficients of −0.293, −0.358, −0.005, and −0.07, respectively as on Table 4. This implies that; as the expenses on these services increase, household expenditure on other goods and services decreases, decreasing household welfare. However, these were statistically significant only for the consultation and laboratory test fees, with p-values of 0.001 and <0.001, respectively.

**Table 4:** Multiple linear regressions for the direct costs associated with average Annual household expenditure.

| Variables | $\beta$ coef | p-value | R squared |
| --- | --- | --- | --- |
| Consultation cost | -0.293 | 0.001 | 0.264 |
| Tests cost | -0.358 | <0.001 |  |
| Drugs cost | 0.189 | 0.028 |  |
| Other drugs cost | 0.327 | <0.001 |  |
| Other treatments cost | -0.005 | 0.95 |  |
| Cost medical equipment | -0.07 | 0.427 |  |

Drugs cost and other drugs cost (auto-medication) had positive regression coefficients of 0.189 and 0.327, respectively, suggesting that the more you spend on drugs and auto-medications, the higher will be your household expenditure, implying better household welfare. These two were, in fact, statistically significant, with p-values of 0.028 and <0.001, respectively. This was controversial to what we expected since spending on these will, in fact, deplete household income, and little will be left to spend for the consumption of other goods and services.

### Effects of indirect cost of kidney failure treatment on the household expenditure of dialysis patients

The regression analysis of each indirect cost component showed that the cost of transportation, caregiver accommodation, informal caregiver cost, and other costs on caregivers had negative regression coefficients of −0.131, −0.055, −0.059, and −0.094, respectively as on table 5. This infers that; as the expenses on these services increase, household expenditure on other goods and services decreases, decreasing household welfare. However, none of these were statistically significant.

**Table 5.** Multiple linear regressions for the indirect cost of kidney failure treatment associated with average annual household expenditure.

| Variables | $\beta$ coef | p-value | R squared |
| --- | --- | --- | --- |
| Transport cost | -0.131 | 0.26 | 0.264 |
| Parking cost | 0.164 | 0.087 |  |
| Accommodation cost | 0.012 | 0.884 |  |
| Caregiver cost | 0.004 | 0.958 |  |
| Caregiver accommodation cost | -0.055 | 0.509 |  |
| Caregiver transport cost | 0.127 | 0.309 |  |
| Informal caregiver cost | -0.059 | 0.483 |  |
| Other caregiver costs | -0.094 | 0.242 |  |

Parking cost, accommodation cost, and caregiver cost caregiver transport cost all had positive regression coefficients of 0.164, 0.012, 0.004, and 0.127, respectively, suggesting that the more you spend on parking, accommodation, caregiver, and caregiver transport, the higher your household expenditure will be implying better household welfare. These parameters may be looked upon as indicators of a high standard of living and, therefore, better household welfare. However, none of them were statistically significant.

## Conclusion

This multi-center study demonstrated that the cost of hemodialysis at the household level in Cameroon is exceptionally high compared with the cost of living and is mainly due to the cost of consultation, drugs, and laboratory investigation. Despite the state subsidy, most patients are at a low socioeconomic level; out-of-pocket expenditure is extremely high and unaffordable for patients and their relatives in the long term. Hemodialysis is an economic burden on households, so strategies to cut these costs should be implemented. The cost of consultation, laboratory tests, other treatment costs (nutrition), and the cost of medical devices had negative correlations. This implies that; increasing the use of these commodities/services at the best minimal cost will decrease the household burden of kidney failure. The indirect cost components showed that the cost of transportation, caregiver accommodation, informal caregiver cost, and other costs on caregivers had a negative correlation; therefore, increasing the use of these commodities/services at best minimal cost will decrease the household burden of kidney failure.

The government should hasten universal health coverage in Cameroon to specifically cover the cost of consultation, laboratory tests, medications and transportation for dialysis patients in these regions.

Kidney disease screening and prevention programs are necessary to reduce the number of persons in need of dialysis and kidney transplants. This remains the only cost-effective and sustainable approach, especially in developing countries like Cameroon. Therefore, the government should implement a policy of annual kidney disease screening especially for people at risk.

## Data Availability

The data underlying the results presented in the study are available on request from Dr Therence Dingana

## Declarations

## Ethics approval and consent to participate

Ethical clearance was obtained from the Institutional Review Board of the Regional Delegation of health for the Northwest Region and the Regional Delegation of health for the Southwest Region. The respondents were adequately informed using the participant’s information section about all the relevant aspects of the study, including its aim, procedures, and anticipated benefits before data were collected. All participants provided verbal or/and written consent to participating in this study. Assent was obtained for children (less than 21years)

## Consent for publication

Not applicable

## Availability of data and materials

The datasets used and/or analysed during the current study are available from the corresponding author on reasonable request.

## Authors’ contributions

TND conceptualised the idea and generated a research proposal from which the study was conducted. TNN, SNN, NCN, LFF, NMA were major contributors in writing the manuscript. FVK analysed and interpreted the patient data regarding the indirect and Direct cost of kidney failure. All authors read and approved the final manuscript.

## Acknowledgements

Our sincere gratitude goes to all the patients at the Dialysis Centres of the Buea and Bamenda Regional Hospitals. We also thank the directors of the above centres for giving us the administrative clearance needed to complete this piece of work.

## REFERENCES

1 WHO. (2021). World Health Statistics.

2 CDC. (2020). About Global NCDs | Division of Global Health Protection | Global Health | CDC. About Global NCDs. https://www.cdc.gov/globalhealth/healthprotection/ncd/global-ncd-overview.html

3 Eknoyan, G., Lameire, N., Barsoum, R., Eckardt, K. U., Levin, A., Levin, N., Locatelli, F., MacLeod, A., Vanholder, R., Walker, R., & Wang, H. (2004). The burden of kidney disease: Improving global outcomes. Kidney International, 66(4), 1310–1314. 10.1111/j.1523-1755.2004.00894.x

4 Himmelfarb, J., Vanholder, R., Mehrotra, R., & Tonelli, M. (2020). The current and future landscape of dialysis. Nature Reviews Nephrology, 16(10), 573–585. 10.1038/s41581-020-0315-4

5 Halle, M. P., M., Kaze, F. F., Fouda, H., Belley, E. P., & Ashuntantang, G. (2017). Cost of care for patients on maintenance haemodialysis in public facilities in Cameroon. African Journal of Nephrology, 20(1), 230–237. 10.21804/20-1-2548

6 Halle, M. P., M., Kaze, F. F., Fouda, H., Belley, E. P., & Ashuntantang, G. (2017). Cost of care for patients on maintenance haemodialysis in public facilities in Cameroon. African Journal of Nephrology, 20(1), 230–237. 10.21804/20-1-2548

7 Allan, J. C., Robert, N. F., Charles, H., Blanche, C., David, G., Charles, H., & Areef Ishani, Kirsten Johansen, Bertram Kasiske, Nancy Kutner, Jiannong Liu, Wendy St Peter, Shu Ding, Haifeng Guo, Allyson Kats, Kenneth Lamb, Shuling, L. A. (2012). US Renal Data System 2012 Annual Data Report - volume2.pdf. University of Minnesota, School of Medicine, USA.

8 Vivekanand, J., Garcia-Garcia, G., & Kunitoshi Iseki, Zuo Li, Saraladevi Naicker, Brett Plattner, Rajiv Saran, Angela Yee-Moon Wang, C.-W. Y. (2013). Chronic kidney disease: global dimension and perspectives. - PubMed - NCBI. Lancet. https://www.ncbi.nlm.nih.gov/pubmed/23727169

9 Vecchi, A. F. De, & M Dratwa, M. E. W. (1999). Healthcare systems and end-stage renal disease (ESRD) therapies--an international review: costs and reimbursement/funding of ESRD therapies. PubMed.

10 Mushi, L., Krohn, M., & Flessa, S. (2015). Cost of dialysis in Tanzania: evidence from the provider’s perspective. Health Economics Review, 5(1). 10.1186/s13561-015-0064-4

11 Hainsworth, T. (2004). The NSF for Renal Services. Part One: Dialysis and transplantation. Nursing Times, 100(5), 28–29.

12 Pockros, B. M., Finch, D. J., & Weiner, D. E. (2021). Dialysis and total health care costs in the united states and worldwide: The financial impact of a single-payer dominant system in the us. Journal of the American Society of Nephrology, 32(9), 2137–2139. 10.1681/ASN.2021010082

13 Xu K, Evans D, Carrin G, A.-R. A. (2008). Designing health financing systems to reduce catastrophic health expenditure. 7. www.who.int/health_financing/pb_2.pdf http://www.who.int/health_financing/documents/pb_e_08_1-cct.pdf

14 NIDDK. Choosing a Treatment for Kidney Failure | NIDDK. In National Institute of Diabetes and Digestive and Kidney Diseases. https://www.niddk.nih.gov/health-information/kidney-disease/kidney-failure/choosing-treatment(2016).

15 WHO. (2005). Empirical evidence on the economic impact of health in the Russian Federation. In Economic impact of health in the Russian Federation Box (Issue 2005, pp. 11– 16).

16 Mushi, L., Marschall, P., & Fleßa, S. The cost of dialysis in low and middle-income countries: A systematic review. BMC Health Services Research, 15(1), 1–10. 10.1186/s12913-015-1166-8 (2015).

17 Datonye, D. A., Pedro, E.-C., & Friday, S. W. A single-center 7-year experience with end-stage renal disease care in Nigeria-. PMC. (2012).

18. seneh, J. B., Kemah, B. L. A., Mabouna, S., Njang, M. E., Ekane, D. S. M., & Agbor, V. N. (2020). Chronic kidney disease in Cameroon: A scoping review. BMC Nephrology, 21(1), 1–11. 10.1186/s12882-020-02072-5

19. Thompson, S., & Wordsworth, S. An annotated cost questionnaire for completion by patients. 1–92. http://dirum.org/instruments/details/28 http://dirum.org/assets/downloads/634432307461163116-An annotated cost questionnaire for completion by patients.pdf. 2001.

20. Adam, A. M. Sample Size Determination in Survey Research. Journal of Scientific Research and Reports, 26(5), 90–97. 10.9734/jsrr/2020/v26i530263 (2020).

21. Collins, A. J., Foley, R. N., Gilbertson, D. T., & Chen, S. C. (2015). United States Renal Data System public health surveillance of chronic kidney disease and end-stage renal disease. In Kidney International Supplements (Vol. 5, Issue 1, pp. 2–7). 10.1038/kisup. 2015.2

22. Valsa J. J., Susan G. J., Joseph R., Arun T. E. T., Philip J. G. Out-of-pocket expenditures, catastrophic household finances, and quality of life among hemodialysis patients in Kerala, India. Hemodial Int. (2022).

23. Okafor, C., & Kankam, C. Future options for the management of chronic kidney disease in Nigeria. In Gender Medicine (Vol. 9, Issue 1 SUPPL.). 10.1016/j.genm.2011.10.002, (2012).

24. Mushi, L., Krohn, M., & Flessa, S. Cost of dialysis in Tanzania: evidence from the provider’s perspective. Health Economics Review, 5(1). 10.1186/s13561-015-0064-4, (2015).

